# COVID-19 herd immunity strategies: walking an elusive and dangerous tightrope

**DOI:** 10.1101/2020.04.29.20082065

**Authors:** Tobias S Brett, Pejman Rohani

## Abstract

The rapid growth in cases of COVID-19 has threatened to overwhelm healthcare systems in multiple countries. In response, severely affected countries have had to consider a range of public health strategies achieved by implementing non-pharmaceutical interventions. Broadly, these strategies have fallen into two categories: i) “mitigation”, which aims to achieve herd immunity by allowing the SARS-CoV-2 virus to spread through the population while mitigating disease burden, and ii) “suppression”, aiming to drastically reduce SARS-CoV-2 transmission rates and halt endogenous transmission in the target population. Using an age-structured transmission model, parameterised to simulate SARS-CoV-2 transmission in the UK, we assessed the prospects of success using both of these approaches. We simulated a range of different non-pharmaceutical intervention scenarios incorporating social distancing applied to differing age groups. We found that it is possible to suppress SARS-CoV-2 transmission if social distancing measures are sustained at a sufficient level for a period of months. Our modelling did not support achieving herd immunity as a practical objective, requiring an unlikely balancing of multiple poorly-defined forces. Specifically, we found that: i) social distancing must initially reduce the transmission rate to within a narrow range, ii) to compensate for susceptible depletion, the extent of social distancing must be vary over time in a precise but unfeasible way, and iii) social distancing must be maintained for a long duration (over 6 months).

## Introduction

Caused by a novel coronavirus, SARS-CoV-2 [1], COVID-19 is an infectious disease capable of severe respiratory illness and death [2]. Since its identification in Wuhan, China, COVID-19 has become an on-going and rapidly expanding global pandemic that is causing substantial mortality and healthcare system strain in multiple countries [3]. While older individuals and those with underlying conditions are most at risk [4], infection has been seen across age-groups [5, 6]. Worryingly, detection of viral loads in the upper respiratory tract suggests potential for pre- and ogliosymptomatic transmission [7, 8, 9]. Due to the absence of a vaccine, current attempts at controlling SARS-CoV-2 spread are focused on social measures that reduce rates of viral transmission: social distancing (a generalised reduction of contact rates between individuals in the population) and self-isolation by symptomatic individuals [10].

Broadly speaking, two distinct approaches to controlling the spread of SARS-CoV-2 have received much attention. The first aims to suppress transmission in the target population (referred hereafter as “suppression”) [10]. Under this objective, control measures reduce viral transmission to such a degree that sustained endogenous transmission is no longer possible. By maintaining control measures in place for a sufficient period of time, the virus will be eliminated in the focal population. The focus will then shift to preventing subsequent reintroduction. The second approach aims to manage or mitigate the negative health impacts (referred hereafter as mitigation) [10]. While suppression aims to ultimately halt local transmission, mitigation aims to reduce the growth rate of the epidemic to ensure disease burden does not overwhelm healthcare systems [3]. In practice, achieving both objectives requires the roll out of the same types of control measures (social distancing and self-isolation), though the necessary intensities and durations vary. At the time of writing, many countries have adopted extensive social distancing measures (including, after some prevarication, the UK [11]) to either mitigate or suppress SARS-CoV-2 spread [3]. However the severe economic costs and acute social pressures associated with social distancing measures inevitably lead to a push for their relaxation [10]. Due to the potentially long wait until a vaccine is available, the UK government has proposed to attempt to achieve herd immunity in the country by allowing a sufficient section of the population to develop natural immunity via exposure to the disease [11].

The consequences of failure to either adequately mitigate or suppress COVID-19 are potentially catastrophic. Due to the many uncertainties surrounding SARS-CoV-2 transmission, authorities are presented with the worst kind of natural experiment. Mathematical modelling is able to assist evaluating the viability of mitigation and suppression as objectives [12], by simulating the impacts of control strategies on viral transmission, hospital burden, fatalities and population-level immunity. We use an age-stratified disease transmission model, taking the UK as an example, to simulate SARS-CoV-2 spread controlled by individual self-isolation and mass social distancing. We simulated various levels of self-isolation effectiveness and three distinct types of social-distancing measures: i) school (including university) closures, ii) work and social place closures, and iii) older individuals social distancing (see Fig 1). We find that suppression is possible with plausible levels of social distancing and self-isolation, however attempting to mitigate COVID-19 long enough to build herd immunity (while maintaining hospital burden at manageable levels) requires fine tuning of control strengths over a long duration-something that will be extremely challenging in practice.

**Figure 1:**
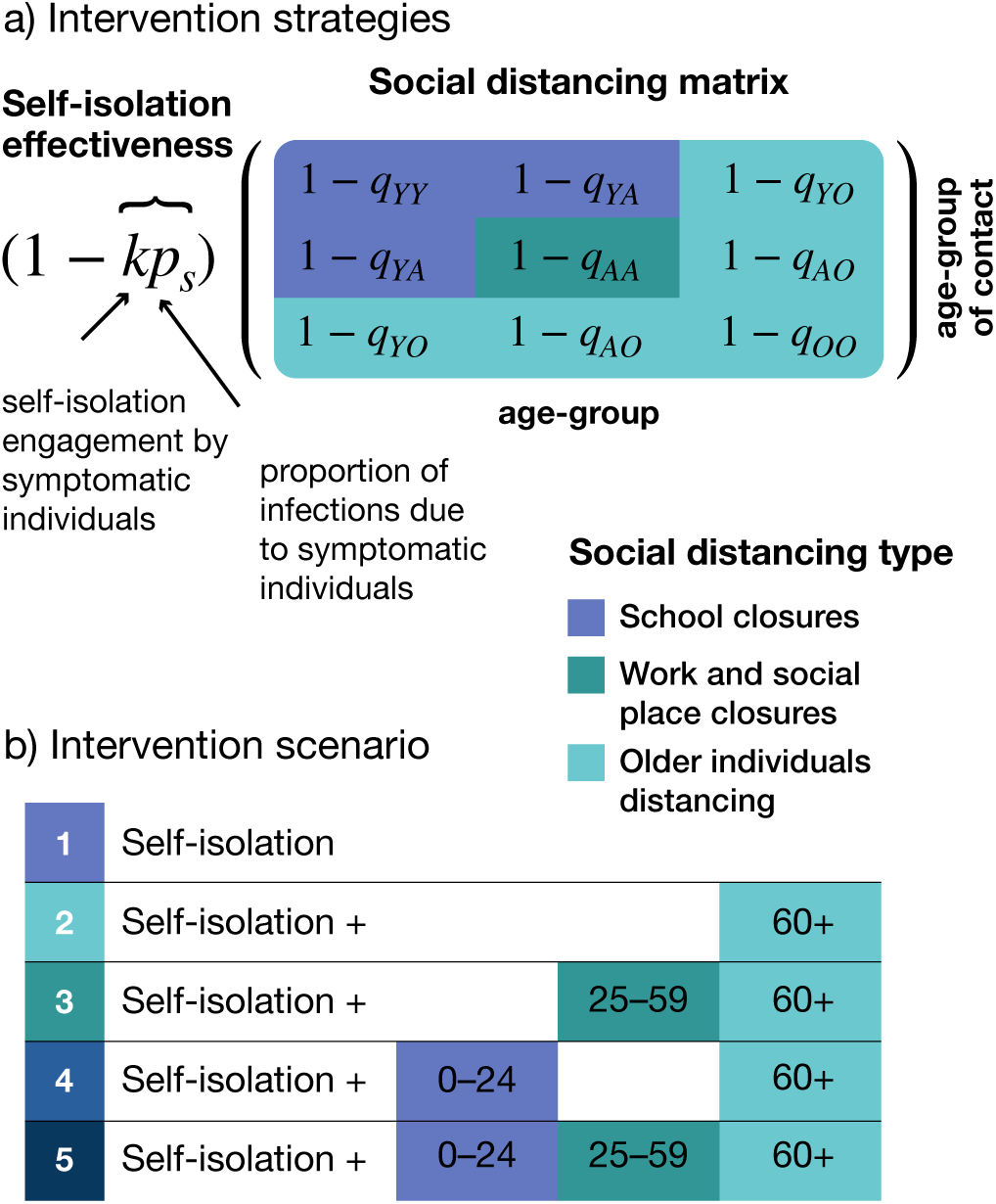
**Modelling the impact of non-pharmaceutical interventions on disease transmission. a) Different social distancing measures (e.g. school closures, work and social place closures, and older individuals distancing) reduce contact rates between individuals in different ways. To reduce the complexity of the model, and to understand their differential impacts, we assume that individuals are only affected by one of these measures, dependent on their age. Individuals aged 0–24 years are affected by school closures (included in this are university closures). School closures were assumed to result in a 70% reduction in contacts among school aged individuals** (*q_YY_*) **and 20 % reduction in their contacts with individuals aged 25–59 years** (*q_YA_*)**. Work and social place closures were assumed to reduce contacts among adults** (*q_AA_*) **by 50%. Finally, older individuals distancing reduced contacts by 60+ aged individuals with 0–24 year-olds by 90%** (*q_YO_*)**, with 25–59 years by 70%** (*q_AO_*) **and among one-another by 50% (***q_OO_***). The effectiveness of symptomatic individuals self-isolating is dependent on two factors: i) the engagement by symptomatic individuals**, *k* **and ii) the proportion of transmission due to individuals who are symptomatic**, *p_s_*. **b) We modelled five distinct combinations of social distancing measures, assuming that older individuals social distancing will always be prioritised**.

## Results

In the absence of any intervention measures, our modelling suggests SARS-CoV-2 will spread extremely rapidly through the UK, with the number of new daily infections exceeding 1 million (Fig 2A). The epidemic would ultimately infect approximately 77% of the population (Fig 2B) and result in around 350 thousand fatalities among individuals aged over 60, and around 60 thousand aged below 60 (Fig 2C).

**Figure 2:**
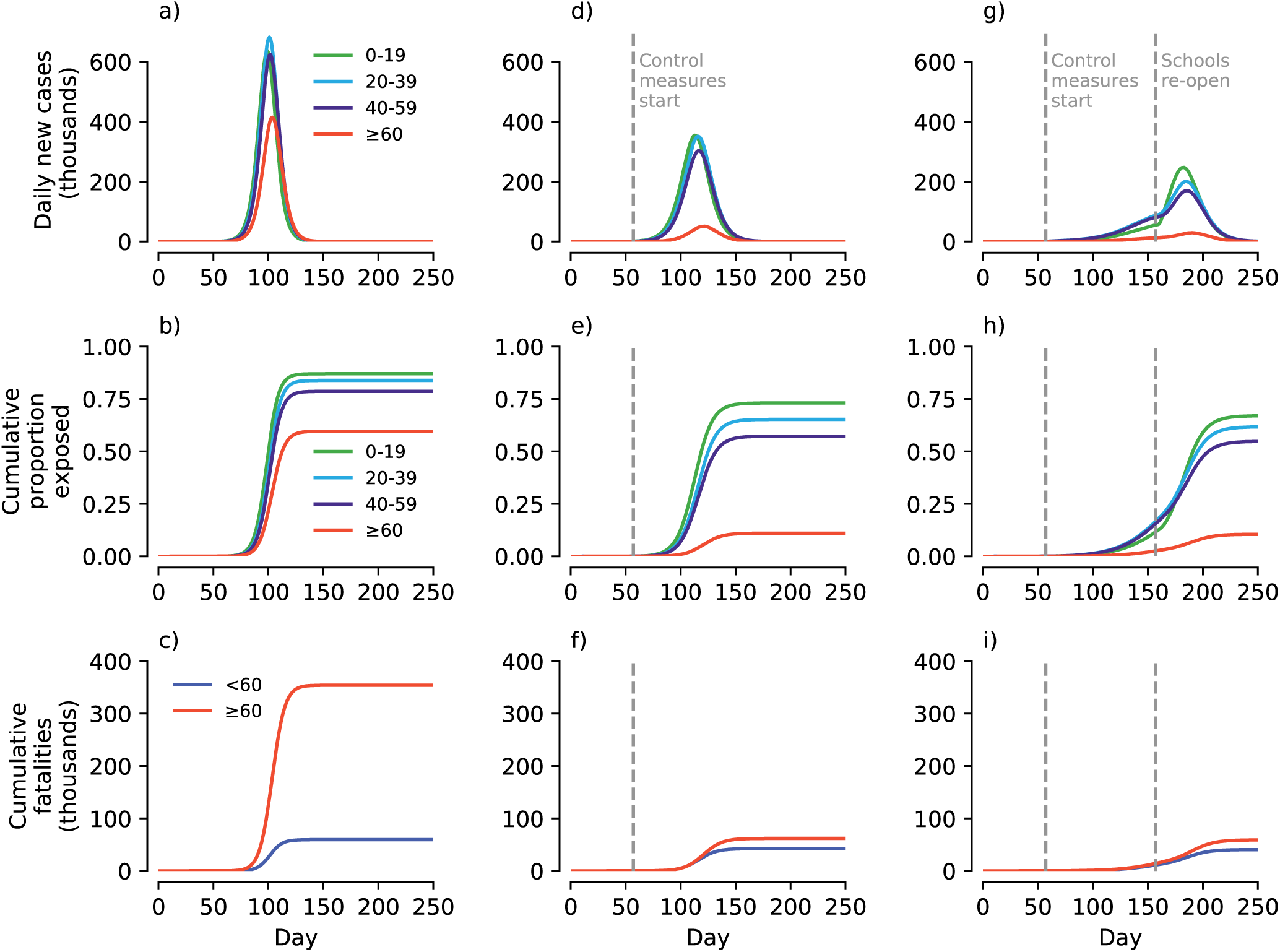
Simulated examples of SARS-CoV-2 spread in the UK using an age-structured SEIR model under different control scenarios. a) Daily new cases assuming no intervention measures enacted. b) Cumulative proportion of the population exposed to SARS-CoV-2 over the course of the epidemic. c) Cumulative fatalities assuming fixed age-specific case fatality rates (see Methods). e-f) Same as panels a-c, but assuming that specific control measures are introduced when daily cases reached 10 thousand (day 57): older individuals social distance and symptomatic individuals self-isolate (at 20% effectiveness). g-i) Same as panels e-f but in addition schools close on day 57. Reopening of schools after 100 days results in a resurgence.

Sustained social-distancing by older individuals (assumed to result in a 90% reduction in contacts with individuals under 25, a 70% reduction with 25–59, and a 50% reduction between one another), and moderately effective self-isolation by symptomatic individuals (at 20% efficacy) results in a shallower epidemic curve (Fig 2D) and a much smaller outbreak size among individuals aged 60+ (Fig 2E). The attendant mortality burden among 60+ individuals is also substantially reduced (to 62 thousand), with a smaller reduction in fatalities in those aged < 60 (to 43 thousand; Fig 2F).

The addition of school (and university) closures, corresponding to a 70% reduction in contacts among school-aged individuals and a 20% reduction with 25–59 year-olds, dramatically reduces the rate of epidemic growth (Fig 2G), although such levels of control are insufficient to suppress the epidemic (i.e. the number of daily cases still rises after implementation). The premature reopening of schools after 100 days (while the virus is still circulating) triggers a second wave of infection, with only a moderately reduced peak in daily new cases, largely eroding any additional gains made [13]. The final proportion of the population exposed (Fig 2H) and the number of fatalities (Fig 2I) are largely unaltered compared to if schools had not been closed (c.f Fig 2E and F).

Our modelling indicates that such control measures can lead to the suppression of COVID-19 in the UK by reducing *R*_0_ < 1 (Fig 3A, B). The effectiveness of self-isolation by symptomatic individuals at suppressing transmission depends on two factors: the proportion of infections due to symptomatic individuals and the self-isolation observance rate (see Fig. 1A). As both of these parameters decrease, the self-isolation efficacy drops, and greater social-distancing measures are necessary to achieve suppression (Fig 3A). At present there is a large uncertainty in the relationship between symptoms and viral shedding [9]. For the social distancing strengths considered, if the proportion due to symptomatic (including mildly symptomatic) individuals is above 14% then, with adequate self-isolation observance, suppression is possible. Given the uncertainty surrounding asymptomatic transmission, the likelihood of successful suppression is greatest if all social distancing measures are enacted (Fig. 3B).

**Figure 3:**
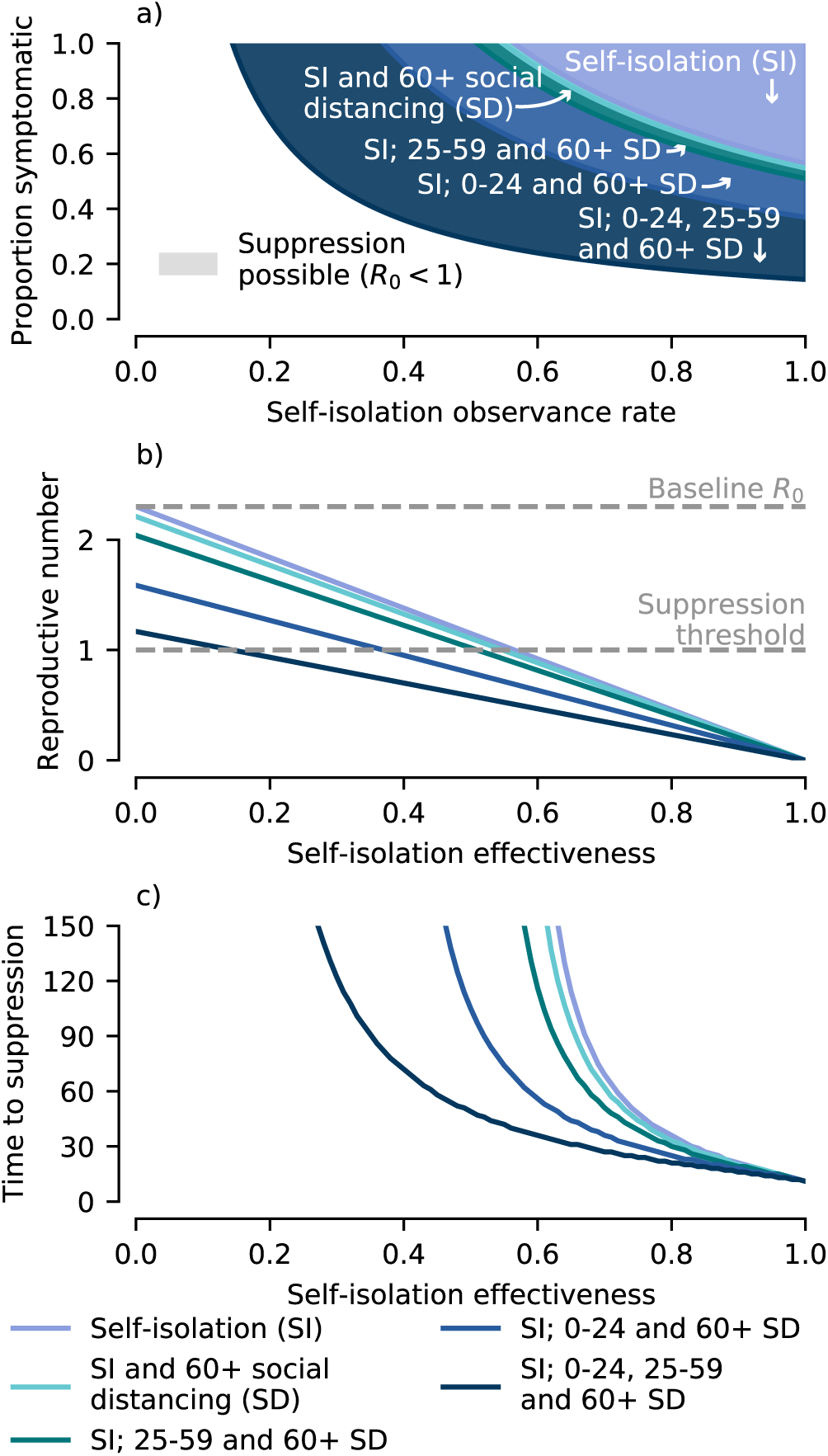
Prospects for disease suppression. Simulations of the age-structured SEIR model performed assuming control measures are initiated when there is a total of 10 thousand infectious individuals in the population. The different control measures and strengths are listed in Fig 1. a) Whether suppression is possible depends on both the self-isolation observance rate and the proportion of infections due to symptomatic individuals. More extensive social distancing measures increase the ranges of these two parameters for which suppression is possible. b) Increases in self-isolation effectiveness drive down the reproductive number, which also depends on the social distancing measures employed. c) The time taken for COVID-19 to be suppressed (modelled as a 100-fold reduction in infectious individuals) depends on the amount the reproductive number is decreased below one.

The time taken for suppression to be achieved (modelled as a 100-fold reduction in infectious individuals) once control measures are implemented is shown in Fig. 3C. If self-isolation effectiveness is high (>70% reduction in transmission) then suppression can be achieved in two months regardless of any additional social distancing measures. There is little additional decrease in the necessary duration of social distancing unless schools and work places are both closed, in which case suppression can be achieved within two months at much lower levels of self-isolation effectiveness ( ≳45%).

If suppression cannot be achieved (due to unfeasibility or lack of political will to reduce transmission sufficiently), then the objective of control measures is mitigation. Social distancing by 60+ aged individuals results in a marked reduction of the final fraction of this age group that are exposed, however, unless both schools and workplaces are closed, additional social distancing measures do not lead to much further reduction (Fig 4A).

**Figure 4:**
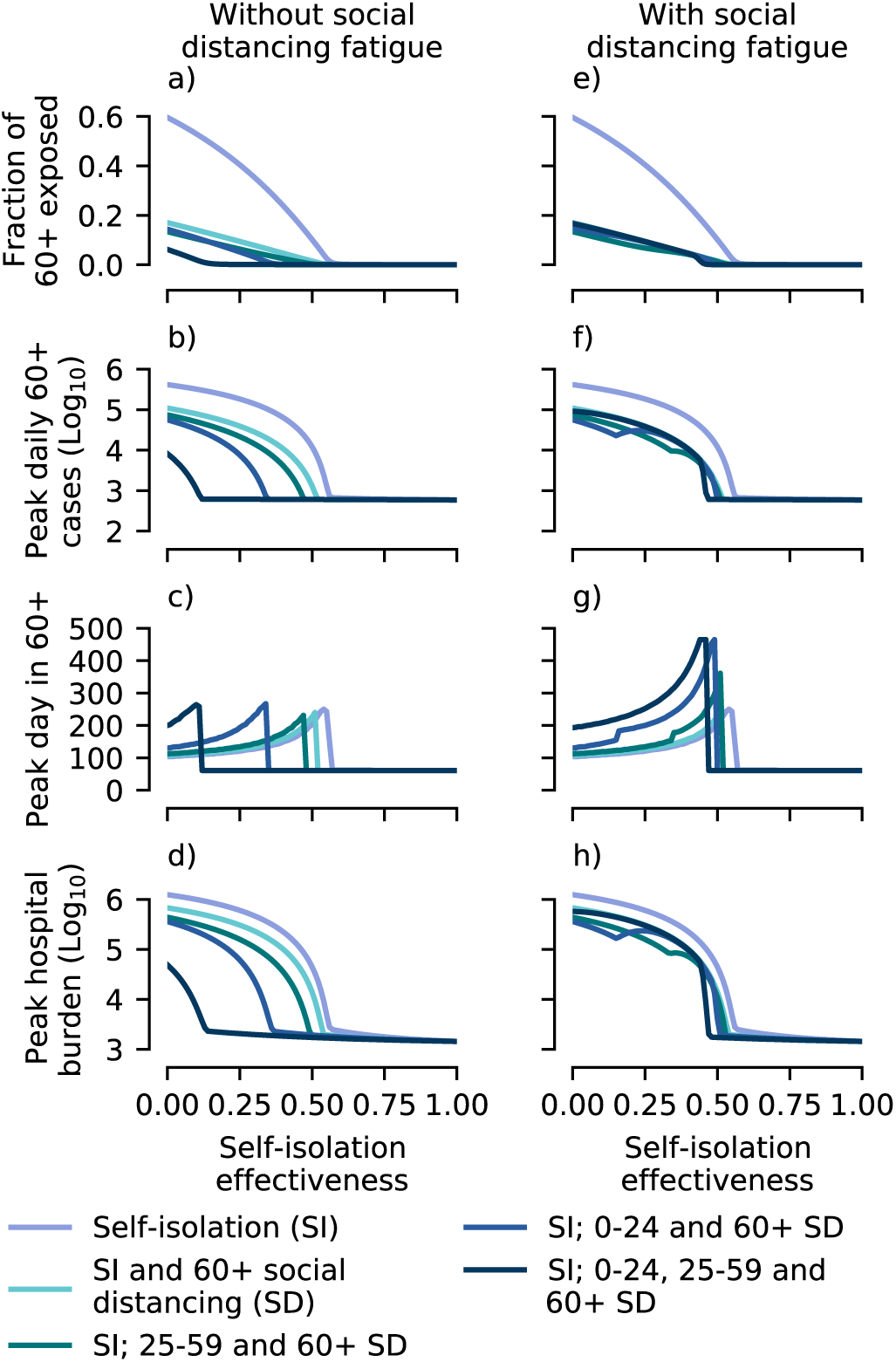
Outcomes of disease mitigation attempts, simulated using the age-structured SEIR model. As in Fig 3, control measures are implemented when there are 10 thousand cases in the population. a) The final fraction of 60+ aged individuals exposed to COVID-19 for each of the control strategies simulated, assuming that social distancing measures can be maintained at the same strength indefinitely (“without fatigue”). b-c) Size (panel b) and timing (panel c) of the peak in daily new cases among 60+ aged individuals. d) Peak hospital burden, assuming age-specific hospitalisation rates (see Methods) and a mean hospital stay of 12.8 days [4]. e-f) Simulation results using the same control strategies as in panels a-d, but assuming that due to fatigue schools and work places closures last 100 days.

These results are also mirrored in the impacts of social distancing on the daily cases in 60+ aged individuals (Fig 4B). Unfortunately, the hospital burden remains high for most intervention strategies, unless self-isolation is very effective (≳ 50%; Fig 4D). Taking around one hundred thousand hospitalised cases to be the upper limit of hospital capacity, we find that there is a relatively small range of parameters where mitigation is successful at preventing hospitals being overwhelmed, but the disease is not also successfully suppressed (Fig 4D, c.f Fig 3B). If social distancing is applied to all age groups, this range is 0–14% self-isolation effectiveness, whereas if just the 60+ age group socially distance the range is 41–54%.

As mentioned previously, if schools and workplaces reopen simultaneously after 100 days (e.g. due to social distancing “fatigue” [10]) and the disease has not been successfully suppressed, then much of the benefit of their closure is lost (Fig 4E-H) due to a resurgent second wave. In this scenario, the principle effect of school and workplace closures is in delaying the peak, buying more time for preparations (Fig 4G).

Assuming the object is mitigation, to prevent a second wave overwhelming the healthcare system, control measures must be relaxed gradually (Fig. 5A). This allows for herd immunity to build up in the population without healthcare systems being overwhelmed. Our modelling suggests this relaxation must take place over a relatively long time span and cannot be linear (a linear relaxation over 300 days results in a peak hospital burden of 99 thousand cases and total fatalities of around 83 thousand, but fails to achieve herd immunity; Fig. 5B).

**Figure 5:**
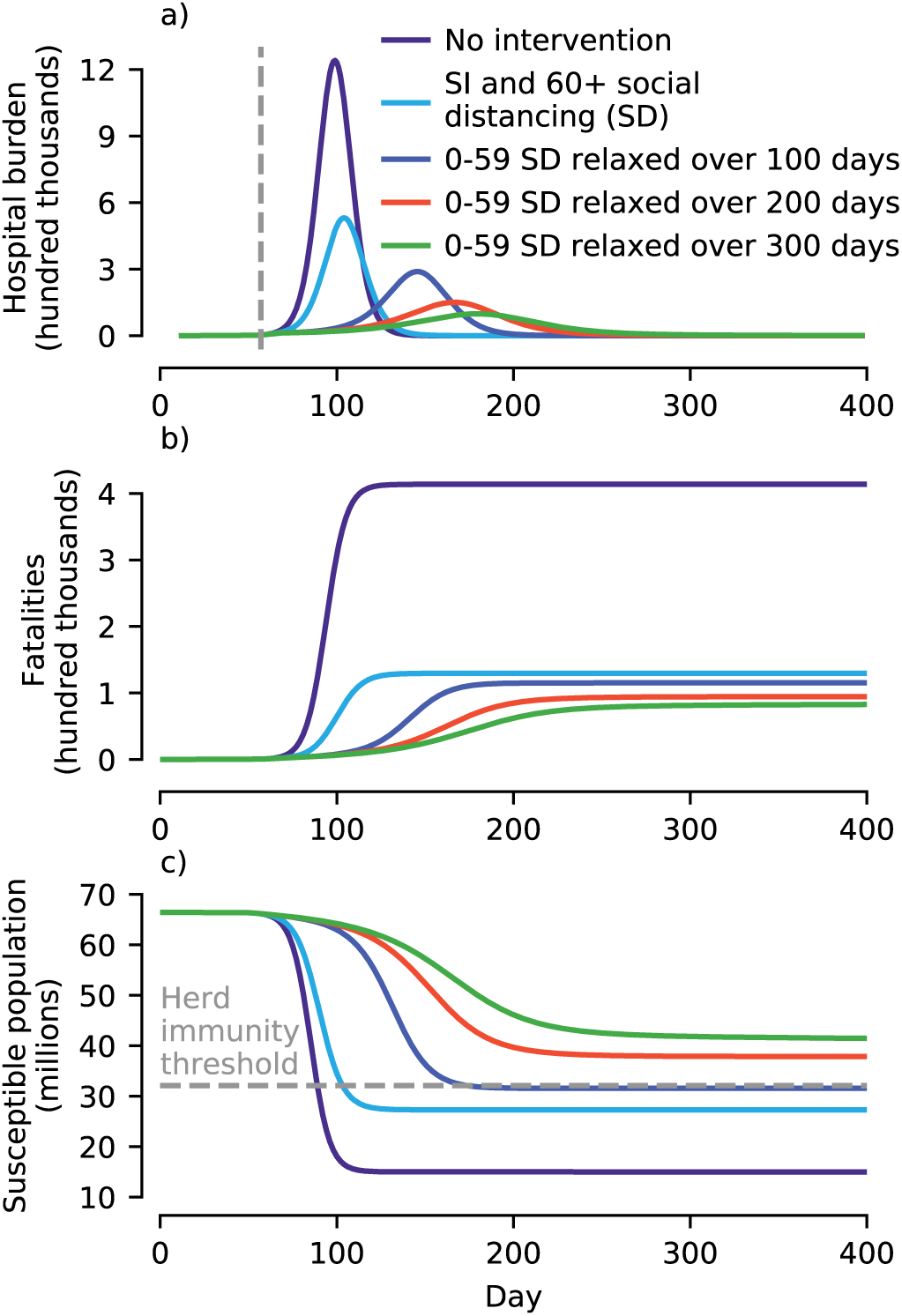
Disease mitigation in the UK with gradual relaxation of controls. Simulated results generated using the age-structured SEIR model. a) Inclusion of social distancing by 0–59 year-olds (e.g. due to school and workplace closures) reduces peak hospital burden. The more gradually social distancing by these age groups is relaxed, the greater the reduction. b) More gradual relaxation in controls results in a slightly smaller final number of fatalities. c) The overall reduction in the susceptible population below the herd immunity threshold is less the longer controls are applied. Simulated results assumed control measures are initiated when there is a total of 10 thousand infectious individuals in the population and that social distancing measures affecting 0–59 year-olds (school and work place closures) are gradually relaxed. For all results shown (apart from where there was no intervention) self-isolation effectiveness was assumed to be maintained at 20%. Similarly, 60+ aged individuals were assumed not to relax their social distancing.

One associated downside to gradually relaxing controls is that the herd immunity achieved is less robust than if no attempts at mitigation were made (Fig. 5C). This is due to the reduced overshoot of the susceptible population below the herd immunity threshold after the epidemic peaks. A smaller subsequent increase in the susceptible population (e.g. due to new births or waning natural immunity) is then required for herd immunity to be lost. The robustness of herd immunity trades off directly with the number of fatalities, as greater susceptible depletion necessitates more infection.

To summarise, aiming to build herd immunity to SARS-CoV-2 in a population while mitigating the burden on hospitals requires initially reducing the reproductive number to ensure available hospital capacity is not exceeded (Fig 6A).

**Figure 6:**
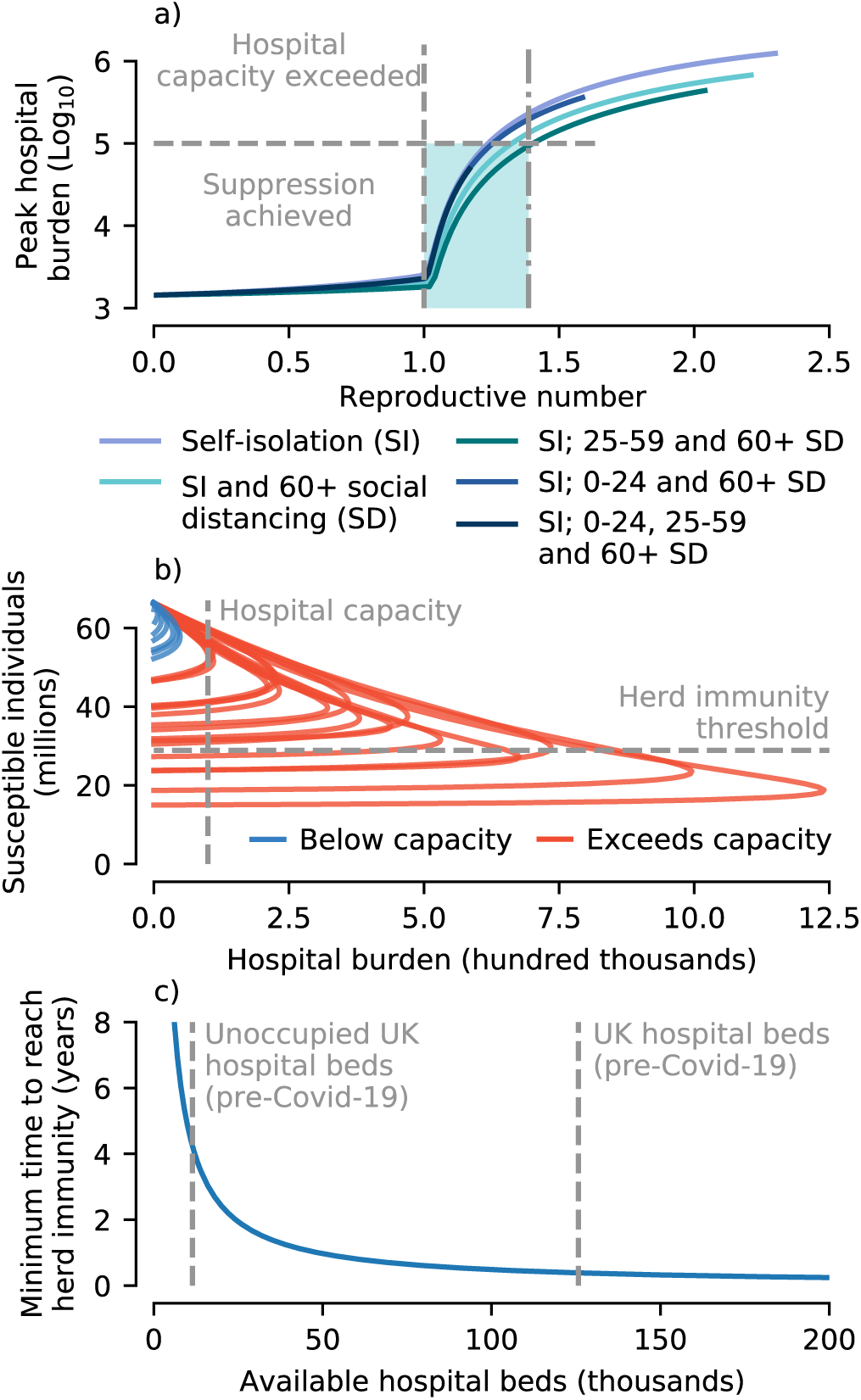
Summary of prospects for achieving herd immunity. a) The peak hospital burden (shown on a log scale) is highly sensitive to the reproductive number. There is a narrow window of reproductive number values (shaded) where either i) the number of COVID-19 cases requiring hospitalisation does not overwhelm hospital capacity (modelled at 100 thousand beds) or ii) circulation is suppressed. b) None of the simulated control scenarios shown in Figs 3 and 4 achieved herd immunity while keeping cases below hospital capacity. c) To achieve herd immunity in the minimum time requires control measures that fix the rate of new hospitalised cases to ensure hospital beds are continually full. This time depends on the available hospital capacity, with the i) total and ii) unoccupied general and acute hospital beds in the UK National Health System pre-COVID-19 indicated.

Without gradual relaxation of social distancing measures, if the epidemic peaks at or below hospital capacity the final outbreak size will be insufficient to achieve herd immunity (Fig 6B). Relaxing social distancing measures linearly also appears insufficient (see Fig 5A)

Even if an optimal strategy for gradually relaxing social distancing to build herd immunity were found, the healthcare system capacity imposes a lower limit of the duration of social distancing required to achieve herd immunity (Fig 6C). Assuming individuals aged 60+ are socially distancing, around 3.7% of cases require hospital treatment. The remaining interventions studied had little further effect on the hospitalisation rate. Assuming a hospital capacity of 100 thousand beds and an average hospital stay of 12.8 days [4], it would take at minimum around 6 months for the UK to achieve herd immunity. We stress that this minimum is unlikely to be found in practice, due to the need for fine tuning social distancing.

## Discussion

Various governments have toyed with the idea of achieving herd immunity through natural infection as a means of ending the long-term threat of COVID-19. This has provoked alarm in sections of the public health community [11,14]. Our work confirms that this alarm is well founded.

Attempting to build up to herd immunity while simultaneously mitigating the impact of COVID-19 on hospital burden is an extremely challenging task. In order to ensure hospital burden in the UK does not exceed 100 thousand beds, *R*_0_ needs to be reduced from its initial value (assumed to be *R*_0_ *=* 2.3) to about 1.4. Suppression is possible if *R*_0_ is reduced below 1. Due to the fine margins (in terms of control effectiveness) between successful disease suppression and overwhelming hospitals, making herd immunity the primary objective (rather than applying maximal social distancing and aiming for suppression) is not supported by our modelling. Put another way, mitigation (via “flattening the curve”) is not a practical objective: if mitigation efforts are sufficient to prevent hospitals from being overwhelmed, only a comparatively small further increase in control measures will drive *R*_0_ below one, and make suppression possible.

In addition to the narrow range of *R*_0_ that must be aimed for, social distancing measures must be gradually relaxed in an highly controlled manner of an long period. Gradually linearly decreasing social distancing was found to be inadequate. Given the estimated proportion of cases that need hospitalisation [15], achieving herd immunity requires finely tuning social distancing over an extended period of time that depends on available hospital capacity. If 100 thousand beds are available for COVID-19 patients (1.52 beds per 1000 people) then at a minimum it will take around 6 months to achieve herd immunity. This is longer than the necessary duration of control measures to achieve disease suppression for almost all control strengths where suppression is possible. We highlight that the different strengths of social distancing were chosen to be plausible values for the UK, erring on the side of caution (underestimating effectiveness). The apparent rapid suppression of SARS-CoV-2 transmission in Wuhan is concordant with greater levels of social distancing than modelled here.

Estimates of hospital burden depend on the average stay of a hospitalised individual. We adopted a value of 12.8 days drawn from a study of cases in Wuhan, China [4]. Other studies estimated the average stay to be 22–24 days [15,16], which almost doubles the hospital burden. Our qualitative conclusions are unaltered by adopting a longer hospital stay, in fact margins for error are further diminished. Re-running the analysis with a hospital stay of 22 days revealed *R*_0_ must be reduced to below 1.3 to avoid exceeding hospital capacity and herd immunity will take over 300 days of social distancing to achieve (results not shown).

As a novel pathogen, there are many epidemiological uncertainties surrounding the spread of SARS-CoV-2. Preliminary evidence suggests that although disease severity is reduced, children at at similar risk of infection as adults [6]. Evidence from Germany suggests a growing role of 15–34 year-olds in driving SARS-CoV-2 transmission [17]. In the absence of clear evidence to the contrary, we therefore made the minimal modelling assumption that infectiousness is not age-dependent. As new evidence comes to light, exploring their impacts on appropriate control measures and strengths is obviously needed. In broad terms, if there is a reduced contribution to transmission from children, then the impact of school closures will be similarly reduced and increased social distancing among adults will be necessary to compensate.

Two other human coronaviruses, HCoV-OC43 and HCoV-HKU1, both cause annual winter-time outbreaks in temperate regions [18], spurring investigation into the effects of seasonality on SARS-CoV-2 circulation [19]. At present, due to the lack of knowledge of how seasonality may impact SARS-CoV-2, we excluded excluded it from our model. Broadly, reductions in transmission due to seasonality will aid in controlling viral spread and mitigating hospital burden. However, additional variability in transmission rates will further complicate attempts relaxing social distancing in a controlled manner to build herd immunity. For instance, if there is a substantial seasonal reduction in transmissibility in summer months then the prospects for temporary mitigation will be enhanced, however if social distancing measures are then halted in response, seasonality may amplify a subsequent resurgent outbreak in winter time [19].

The estimates of hospital burden and fatalities were calculated using results from a study on cases in Wuhan, China [15]. For this study we took the point estimates, however these had uncertainties associated with them and they are unlikely to be the same across regions. There is an obvious feedback between fatality rates and healthcare system burden, however it is unclear the extent to which the Wuhan healthcare system (used in estimation) was overwhelmed. We therefore assumed the fatality rates fixed regardless of hospital burden. For these reasons we have avoided attaching confidence intervals to estimates of fatalities, and they should be interpreted as plausible projections and not predictions.

Similarly, [15] assumed that the fraction of cases hospitalised in Wuhan was equal to the rate of severe disease. To the best of our knowledge, no information on what fraction of these cases required an intensive treatment unit (ITU) bed or use of ventilator is available. We therefore adopted 100 thousand contemporaneous hospitalisations as an indicative threshold level of hospital burden. Prompt publication of the proportions of UK COVID-19 cases requiring hospitalisation and intensive treatment (and also hospital and ITU capacities) will enable modellers to more accurately model/gauge healthcare system burden.

A major unknown remains the nature, duration and effectiveness of natural immunity. Here, we made the pragmatic assumption that, over the time scales under consideration, infection confers perfect long-lasting immunity (the best case scenario for mitigation strategies). If immunity is not perfect, and there is a moderate to high chance of reinfection, then prospects for achieving herd immunity via natural infection are slim [19]. To shed light on the kinetics of immunity, mass longitudinal antibody testing is necessary. This would both permit the identification of previously infected individuals, and provide information regarding immunity through time [20]. We submit models such as the one explored here provide a powerful means of integrating parallel serological and epidemiological data streams to quantify population-level immunity. Further, such models can be central to the development of efficient age-stratified serological testing schemes.

Finally, we stress that our study only explored the epidemiological impacts of non-pharmaceutical interventions (social distancing and self-isolation). Ultimately, any comprehensive public health policy needs to take into account the concomitant and wide-ranging societal and economic consequences of control measures.

## Methods

### Model

We used a deterministic age-structured SEIR transmission model to simulate COVID-19 transmission in the United Kingdom. Contact rates *c_i,j_*, the number of daily contacts an individual of age *i* makes with individuals of age *j*, were taken from the POLYMOD study for the UK [21] corrected for reciprocity [22]. The simulated age groups were matched to those of the POLYMOD study: 14 5-year increments from 0 to 69 and then 70+. Age-stratified population sizes (*N_j_*) were taken from 2018 UK demographic data.

The mean latent and infectious periods were set to 1*/ρ* = 3 and 1/*γ* = 3 days respectively, consistent with various estimates of the serial interval [23, 24] and incubation period [4, 25, 26], assuming infectiousness starts 1–2 days before symptoms develop.

Both latent and infectious periods were assumed to be gamma distributed and modelled using the method of stages [27, 28, 29], by dividing the exposed (*E_i_*) and infectious (*I_i_*) compartments for each age class into 4 sub-compartments, 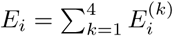 and 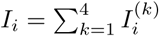. where the superscript labels the sub-compartment. The transmission dynamics for the age classes were governed by a system of ordinary differential equations,

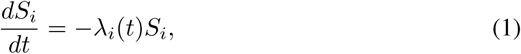

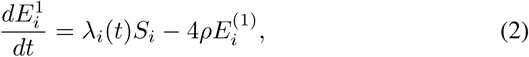

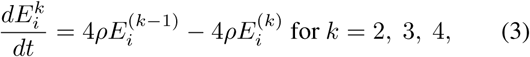

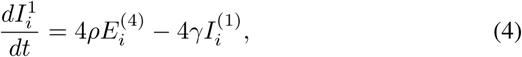

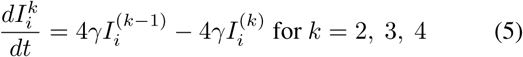

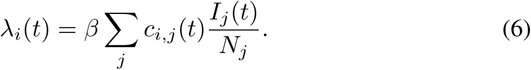

The transmission rate *β* was tuned using the next-generation matrix [30] to give a value of *R*_0_ = 2.3, consistent with estimates [23, 24]. Simulations were initialised with one initial introduction in a fully susceptible population (*S_i_* = *N_i_*). The resulting doubling time was observed to be about 3 days, broadly consistent with early observations from the UK.

Model is set up in such a way that *c_i,j_* and can be manipulated to account for various control measures (see next section).

### Modelling non-pharmaceutical interventions

Two types of on-pharmaceutical intervention were modelled: i) self-isolation by symptomatic infectious individuals and ii) mass social distancing by differing age groups. The effectiveness of self-isolation of symptomatic individuals is dependent on the product of two factors: i) the proportion of infections that occur due to symptomatic individuals, *p_s_* and ii) the observance rate of social-isolation among symptomatic individuals, *k*. The fractional reduction of contacts between age classes *i* and *j* due to social distancing is given by *q_i,j_*.

Both of these interventions take the form of modifications to the contact matrix between infectious and susceptible individuals,

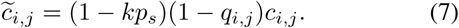

This expression for 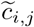is inserted in place of *c_i_,_j_* in Eq. 6.

Age classes in the model are divided into whether they are young (*Y*; corresponding to 0–24 year olds and age groups *i* = 1 to 4), adults (*A*; 25–59 year olds, age groups *i* = 5 to 11) and older (*O*; 60+ year olds, age groups *i* = 12 to 15). The reduction in contacts due to social distancing, *q_i,j_*, is then determined by which of these three categories the contacter and contactee fall into, given by the block matrix

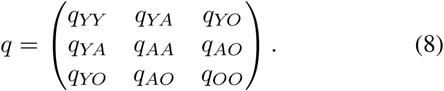

We assume school closures reduce contact rates between young individuals by a factor of *q_YY_* = 0.7 and between young people and adults by *q_YA_* = 0.2. Social distancing among adults (e.g. due to workplace closures and reduction in social events) was modelled as a reduction of *q_AA_* = 0.5. Social distancing of older individuals was represented by *q_YO_* = 0.9, *q_AO_* = 0.7, *q_OO_* = 0.5. For simulations with social distancing fatigue, *q_YY_*, *q_YA_* and *q_AA_* were modelled as linearly decreasing from these initial values to 0 over the periods indicated in Fig 5.

### Estimating hospital burden and case fatalities

Age-specific hospitalisation and fatality rates were taken from point estimates calculated in a study on cases in Wuhan, China [15]. Due to differences in the final age group of our model (70+) and those of the Wuhan study (70–79 and 80+), the hospitalization and fatality rates for 70+ individuals were calculated by summing the estimated 70–79 and 80+ rates weighted by their relative population sizes. Based on results from a separate study on cases in Wuhan, China, we assumed the average duration of hospitalisation with COVID-19 was 12.8 days [4].

Pre-COVID-19 hospital beds and occupancy rates in the UK National Health Systems (NHS) were taken from the most recent (autumn 2019) published numbers for Northern Ireland, Wales, Scotland, and England.

### Code availability

All code and data used in this study are available at https://github.com/tsbrett/COVID-19_herd_immunity/releases/tag/v1.0.

## Acknowledgments

This work is funded by the National Institute of Health through a MIDAS (Models of infectious disease agent study) grant 5R01GM123007.

## References

[1] Zhu N, Zhang D, Wang W, Li X, Yang B, Song J, et al. A novel coronavirus from patients with pneumonia in China, 2019. New England Journal of Medicine. 2020;.

[2] Huang C, Wang Y, Li X, Ren L, Zhao J, Hu Y, et al. Clinical features of patients infected with 2019 novel coronavirus in Wuhan, China. The Lancet. 2020;395(10223):497–506.

[3] Bedford J, Enria D, Giesecke J, Heymann DL, Ihekweazu C, Kobinger G, et al. COVID-19: towards controlling of a pandemic. The Lancet. 2020;.

[4] Guan Wj, Ni Zy, Hu Y, Liang Wh, Ou Cq, He Jx, et al. Clinical characteristics of coronavirus disease 2019 in China. New England Journal of Medicine. 2020;.

[5] Liu W, Zhang Q, Chen J, Xiang R, Song H, Shu S, et al. Detection of Covid-19 in children in early January 2020 in Wuhan, China. New England Journal of Medicine. 2020;.

[6] Bi Q, Wu Y, Mei S, Ye C, Zou X, Zhang Z, et al. Epidemiology and Transmission of COVID-19 in Shenzhen China: Analysis of 391 cases and 1,286 of their close contacts. medRxiv. 2020;.

[7] Zou L, Ruan F, Huang M, Liang L, Huang H, Hong Z, et al. SARS-CoV-2 viral load in upper respiratory specimens of infected patients. New England Journal of Medicine. 2020;382(12):1177–1179.

[8] Wei WE, Li Z, Chiew CJ, Yong SE, Toh MP, Lee VJ. Presymptomatic Transmission of SARS-CoV-2—Singapore, January 23-March 16, 2020. Morbidity and Mortality Weekly Report. 2020;69(14):411.

[9] Wölfel R, Corman VM, Guggemos W, Seilmaier M, Zange S, Müller MA, et al. Virological assessment of hospitalized patients with COVID-2019. Nature. 2020;p. 1–10.

[10] Anderson RM, Heesterbeek H, Klinkenberg D, Hollingsworth TD. How will country-based mitigation measures influence the course of the COVID-19 epidemic? The Lancet. 2020;395(10228):931–934.

[11] Hunter DJ. Covid-19 and the Stiff Upper Lip—The Pandemic Response in the United Kingdom. New England Journal of Medicine. 2020;.

[12] Anderson RM, Anderson B, May RM. Infectious diseases of humans: dynamics and control. Oxford university press; 1992.

[13] Hollingsworth TD, Klinkenberg D, Heesterbeek H, Anderson RM. Mitigation strategies for pandemic influenza A: balancing conflicting policy objectives. PLoS computational biology. 2011;7(2).

[14] Horton R. Offline: COVID-19—a reckoning. Lancet (London, England). 2020;395(10228):935.

[15] Verity R, Okell LC, Dorigatti I, Winskill P, Whittaker C, Imai N, et al. Estimates of the severity of coronavirus disease 2019: a model-based analysis. The Lancet Infectious Diseases. 2020 03;.

[16] Zhou F, Yu T, Du R, Fan G, Liu Y, Liu Z, et al. Clinical course and risk factors for mortality of adult inpatients with COVID-19 in Wuhan, China: a retrospective cohort study. The Lancet. 2020;.

[17] Goldstein E, Lipsitch M. Temporal rise in the proportion of both younger adults and older adolescents among COVID-19 cases in Germany: evidence of lesser adherence to social distancing practices? medRxiv. 2020;.

[18] Killerby ME, Biggs HM, Haynes A, Dahl RM, Mustaquim D, Gerber SI, et al. Human coronavirus circulation in the United States 2014–2017. Journal of Clinical Virology. 2018;101:52–56.

[19] Kissler SM, Tedijanto C, Goldstein E, Grad YH, Lipsitch M. Projecting the transmission dynamics of SARS-CoV-2 through the postpandemic period. Science. 2020;.

[20] Antia A, Ahmed H, Handel A, Carlson NE, Amanna IJ, Antia R, et al. Heterogeneity and longevity of antibody memory to viruses and vaccines. PLoS biology. 2018;16(8):e2006601.

[21] Mossong J, Hens N, Jit M, Beutels P, Auranen K, Mikolajczyk R, et al. Social contacts and mixing patterns relevant to the spread of infectious diseases. PLoS medicine. 2008;5(3).

[22] Riolo MA, Rohani P. Combating pertussis resurgence: One booster vaccination schedule does not fit all. Proceedings of the National Academy of Sciences. 2015;112(5):E472-E477.

[23] Li Q, Guan X, Wu P, Wang X, Zhou L, Tong Y, et al. Early transmission dynamics in Wuhan, China, of novel coronavirus-infected pneumonia. New England Journal of Medicine. 2020;.

[24] Zhang J, Litvinova M, Wang W, Wang Y, Deng X, Chen X, et al. Evolving epidemiology and transmission dynamics of coronavirus disease 2019 outside Hubei province, China: a descriptive and modelling study. The Lancet Infectious Diseases. 2020;.

[25] Linton NM, Kobayashi T, Yang Y, Hayashi K, Akhmetzhanov AR, Jung Sm, et al. Incubation period and other epidemiological characteristics of 2019 novel coronavirus infections with right truncation: a statistical analysis of publicly available case data. Journal of clinical medicine. 2020;9(2):538.

[26] Lauer SA, Grantz KH, Bi Q, Jones FK, Zheng Q, Meredith HR, et al. The incubation period of coronavirus disease 2019 (COVID-19) from publicly reported confirmed cases: estimation and application. Annals of internal medicine. 2020;.

[27] Lloyd AL. Realistic distributions of infectious periods in epidemic models: changing patterns of persistence and dynamics. Theoretical population biology. 2001;60(1):59–71.

[28] Wearing H, Rohani P, Keeling M. Appropriate models for the management of infectious diseases. PLoS Medicine. 2005;2(7):621.

[29] Keeling MJ, Rohani P. Modeling Infectious Diseases in Humans and Animals. Princeton University Press; 2008.

[30] Van den Driessche P, Watmough J. Reproduction numbers and sub-threshold endemic equilibria for compartmental models of disease transmission. Mathematical biosciences. 2002;180(1–2):29–48.

